# Cross-Sectional Study of University students’ attitudes to ‘on campus’ delivery of COVID-19 vaccines and future-proofing MenACWY and MMR vaccination rates by adopting COVID-19 vaccine roll-out strategies

**DOI:** 10.1101/2022.02.07.22270394

**Authors:** Adam Webb, Mayuri Gogoi, Sarah Weidman, Katherine Woolf, Maria Zavala, Shamez N Ladhani, Manish Pareek, Lieve Gies, Christopher D Bayliss

**Affiliations:** Department of Genetics and Genome Biology, University of Leicester, UK; Department of Respiratory Sciences, University of Leicester, UK; University College London Medical School, London, UK; Immunisation and Countermeasures Division, Public Health England Colindale, London, UK; Department of Infection and HIV Medicine, University Hospitals of Leicester NHS Trust, UK; School of Media, Communication and Sociology, University of Leicester, UK

## Abstract

**Background:** University students are a critical group for vaccination programmes against COVID-19, meningococcal disease (MenACWY), and measles, mumps and rubella (MMR). We aimed to evaluate risk factors for vaccine hesitancy (refusal or intention to refuse a vaccine) and views of university students about on-campus vaccine delivery.

**Methods:** Cross-sectional anonymous online questionnaire study of undergraduate students at a British university in June 2021. Chi-squared, Fisher’s exact, univariate and multivariate tests were applied to detect associations.

**Results:** Complete data were obtained from 827 participants (7.6% response-rate). Two-thirds (64%; 527/827) reported having been vaccinated against COVID-19 and a further 23% (194/827) agreed to be vaccinated. Other responses were either unclear (66) or indicated an intention to refuse vaccination (40). Hesitancy for COVID-19 vaccines was 5% (40/761). COVID-19 vaccine hesitancy was associated with black ethnicity (aOR, 7.01, 95% CI, 1.8-27.3) and concerns about vaccine side-effects (aOR, 1.72; 95% CI, 1.23-2.39). Lower levels of vaccine hesitancy were detected amongst students living in private accommodation (aOR, 0.13; 95% CI, 0.04-0.38) compared to those living at home. Uncertainty about their personal vaccine status was frequently observed for MMR (11%) and MenACWY (26%) vaccines. Campus-associated COVID-19 vaccine campaigns were definitely (45%) or somewhat (16%) favoured by UK-based students and more so among UK-based international students (62% and 12%, respectively).

**Conclusions:** Vaccine hesitancy among students of black ethnicity and those living at home requires further exploration because attitudes in these groups may affect COVID-19 vaccine uptake. High levels of uncertainty among students about their MMR and MenACWY vaccine status are also a concern for the effectiveness of these vaccine programmes. This issue could be tackled by extending the capabilities of digital platforms for accessing vaccine information, such as the NHSapp in the UK. Sector-wide implementation of on-campus vaccine delivery may also improve vaccine uptake, especially for international students.

## Introduction

Young people are an important risk group for vaccination programmes due to their high mobility, inexperience of accessing medical systems and relatively higher levels of vaccine hesitancy compared to older populations [1,2]. Within this group, university students are particularly at risk of contracting and transmitting infectious diseases because of their high levels of transmission-associated behaviours at university and mixing of geographically-diverse intakes [3-5]. These risks have been exemplified by recent outbreaks of COVID-19 on United Kingdom (UK) university campuses as students returned to campus-based activities after initial lockdown [6]. Facilitating access of university students to vaccines is a key mechanism for enhancing vaccine uptake and preventing infectious diseases outbreaks while minimising the need for highly restrictive measures such as lockdown measures, including social distancing and online learning.

Multiple previous studies have been conducted to determine the reasons for COVID-19 vaccine hesitancy in the general population. Vaccine hesitancy or acceptance has been assessed by a range of measures including use of the SAGE guidelines, Likert scaled-acceptance questions, attitudinal measures and actual uptake or intention to uptake (as utilised herein) [7-10]. A recent meta-analysis of vaccine acceptance in higher income countries reported vaccine hesitancy rates of at least 30% in half the studies (n=97) with lower socio-economic status being the most impactful contributory factor in lower-middle income countries/regions and perceived vaccine safety in more affluent countries/regions [7]. Common demographics for COVID-19 vaccine hesitancy in the reported literature include females, younger age groups, being from a minority ethnic group and lower education or income levels. Studies of student populations have yielded a range of findings. Factors associated with higher vaccine acceptance are knowledge of COVID-19 vaccines, trust in authorities and high perceived vaccine effectiveness [9-14] while perceived accessibility barriers (physical or financial) have been associated with vaccine hesitancy [10, 14]. A potentially important issue is whether concerns about the rare but serious side-effects of the licensed COVID-19 vaccines might have affected uptake among students [15, 16].

Prior to the COVID-19 pandemic, a major concern for student populations was the prevention of cases and outbreaks of meningococcal disease, measles and mumps. Rising levels of infections due to a MenW cc11-lineage strains led to introduction of the MenACWY vaccine into the UK school-age vaccination programme and new university entrants from August 2015 [17, 18]. Outbreaks of mumps among students were also observed in 2019 leading to student-focussed information campaigns to encourage uptake of the MMR vaccine [19]. The MenACWY and MMR vaccines are currently offered free-of-charge to all university students in the UK, including overseas students. National lockdowns to contain the rapid spread of SARS-CoV-2 in 2020 led to major reductions in cases of meningococcal disease, measles and mumps but there is now a concern that ending lockdowns and increased social mixing could lead to rises in these serious vaccine-preventable diseases [20, 21]. These effects may be compounded by disruption of school-based immunisation programmes during the pandemic, which may have resulted in a serious risk of long-term weakening of individual and herd (population) protection.

Studies prior to the COVID-19 pandemic reported uptake rates of the MenACWY vaccines among students at 68-71% [8, 18, 22]. In general, students are expected to obtain their vaccines prior to arrival at university. However, uptake can be enhanced by ‘on campus’ vaccine campaigns as exemplified by the University of Nottingham’s highly effective delivery of the MenACWY vaccine for incoming university entrants [18]. Vaccine hesitancy has been examined for the MenACWY vaccine. Blagden et al. [8] reported that vaccine uptake was strongly associated with a high perceived effectiveness of the vaccine but did not find any barriers, such as vaccine side-effects or inconvenience. A meta-study by Wishnant *et al*. [23] found that the only factors strongly associated with uptake of meningococcal vaccines among students were perceived risks of contracting meningococcal disease and the severity of meningococcal infections. Overall, these studies indicate that vaccine hesitancy is not a major barrier to meningococcal vaccine uptake but are equivocal about how vaccine uptake can be increased.

To evaluate the barriers to uptake of vaccines among students and to inform university vaccination policies, we assessed the attitudes, knowledge, perceived vaccine status and willingness for uptake of COVID-19, MMR and MenACWY vaccines among university students during the roll-out of COVID-19 vaccines to 18-year olds in the UK.

## Material and methods

### Ethics

Ethical approval was given by the University of Leicester (UoL) Medicine and Biological Sciences Research Ethics Committee (reference number 29522). All study participants provided written informed consent.

### Context of questionnaire delivery and derivation

The questionnaire was emailed to students on three occasions between the 1^st^ and 21^st^ of June 2021. Access to COVID-19 vaccines in the UK was extended to the 25-29, 23-24, 21-22 and 18-20 age brackets on the 7^th^, 15^th^, 16^th^ and 17^th^ June 2021, respectively (https://www.england.nhs.uk/2021/06/21-and-22-year-olds-to-be-offered-covid-19-jab-from-today/ and https://www.england.nhs.uk/2021/06/nhs-invites-all-adults-to-get-a-covid-jab-in-final-push/). Prior to these dates, only healthcare workers and medical students as well as individuals in vulnerable categories were eligible for COVID-19 vaccines <30 year-olds. At the time the questionnaire was designed, we assumed most students would be unvaccinated when they completed the questionnaire.

### Questionnaire delivery, structure and content

The questionnaire was administered via Online Surveys. Between 1^st^ and 21st June 2021, the research team sent an invitation email, and two reminder emails, to all 10,869 campus-based University of Leicester undergraduate students (S1 Figure). Each invitation email contained a unique link to the questionnaire that could not be re-used. Completed questionnaires were de-identified by automatic assignment of another unique identifier by the software, thereby uncoupling the questionnaires from the original email address. The initial email included an invitation to voluntarily participate in follow-up interviews and a prize draw with five prizes of £200 being offered and subsequently delivered.

The questionnaire consisted of a participant information sheet followed by three consent questions. Other parts of the questionnaire were only accessible if consent to all three consent questions was provided. The questionnaire consisted of 29 questions split into four sections: demographics; vaccines; experiences of COVID-19 disease; and other pandemic experiences (e.g. harassment). Questions were multiple choice or scaled answers with one free text box and three questions with answer-dependent questions (S1 Table). Questions 2, 6, 7, 10, 12, 17, 19, 20 and 26 were identical to or modifications of questions utilised in UK-REACH questionnaire 2_ver_1.2 (23 Mar 2021). Questions 13-16 were written by the authors and piloted with University of Leicester students prior to the pandemic as part of another study [22]. Questions 3-5, 8, 9, 11, 21-25, 27-29 were written by the authors for this study. Question 26 is the self-determination scale. Question 17 utilised four statements from the VAX scale of Martin and Petrie [24]. A VAX score was derived for each participant by reversing the scores for statements 17.2, 17.3 and 17.5 followed by re-scaling the sum of all four scores on a 0 to 1 scale that represents maximum to minimum hesitancy, respectively.

### Definitions of primary endpoints

Primary endpoints in our analyses were vaccine hesitancy, VAX scores and willingness for on-campus vaccination programmes. Vaccine hesitancy was defined as providing a response to question 10 that included the phrase ‘have decided not to have the vaccine’. Vaccine willing students were those whose response included either ‘I have already had’ or ‘intend to have the vaccine’. VAX scores have been utilised as predictors of vaccine hesitancy [24]. VAX scores were derived for all students and analysed for differences between ethnic groups and term time residence locations as an alternate measure of vaccine hesitancy. The potential utilisation of on-campus vaccination programmes was defined based on responses to question 15 split between those in favour (definitely increase, somewhat increase) and those who were ambivalent (neither, somewhat decrease, definitely decrease).

### Statistical analysis

De-identified survey responses were analysed using R version 4.0.3 with the tidyverse (data handling), jsonlite 1.7.1 (data extraction), ggplot2 3.32 (general graphing), gtsummary 1.4.2 (tabulation), UpSetR 1.4.0 (graphing of sets) and likert 1.3.5 (graphing of likert-style responses) packages [25-31]. In order to determine if the demographics of our study participants were similar to those of other UK universities and for weighting of the multivariable analyses, we obtained demographic data from HESA (Higher Education Statistics Agency; https://hesa.ac.uk). Similarly, we compared vaccination rates in our study with local, regional and national vaccination rates obtained from the UK Coronavirus Dashboard (https://coronavirus.data.gov.uk).

Univariable analyses were performed on unweighted survey results using chi-squared, Fisher’s exact or Wilcoxon’s rank sum tests. False discovery rate (FDR) correction was used to adjust p-values for multiple testing. COVID-19 vaccine hesitancy and preference for on-campus vaccinations (COVID-19 and MMR) were dichotomized and used as dependent variables. Multivariable analysis was performed using logistic regression on weighted survey results, with vaccine hesitancy and preference for on-campus vaccinations (COVID-19 and MMR) as dependent variables. Predictors included gender, ethnic group, age group, course studied, year of study, experience of harassment, experience of COVID-19 related death, concern over side-effects from the Oxford/AstraZeneca vaccine, concern over hospitalization from COVID-19, concern over spreading COVID-19, home area (local, national, international), residence while studying (home, halls of residence, private accommodation, other) and a psychometric score on self-determination/fatalism. Home area was derived using information on post-codes and international status with students being classified as local if they came from either Leicester or the wider county of Leicestershire, ‘national’ for students from the rest of the UK and ‘international’ for students ordinarily living overseas. Experiences of COVID-19 related deaths were classified into a Yes or No category according to responses to question 21 (S1 Table) with the Yes responses including family member(s), friend(s) or someone else. Survey data were weighted using the raking method in the survey package 4.0 for R based on national student distributions for ethnic group (White, Asian, Black and Other) and gender (S2, S3 and S4 Tables). Constraints in the national data made it necessary to remove students of unknown gender (n = 6).

Statistical differences between the distributions of VAX scores for different groups were determined using a Games-Howell pairwise test with FDR correction.

## Results

### Response rate, sample characteristics and COVID-19 vaccination uptake

In June 2021, all University of Leicester (UoL) undergraduate students were invited to participate in a study of the uptake and attitudes to COVID-19 vaccines. Complete answers were provided by 7.6% (827/10,869) of participants. Respondents were young (94% 18-25 year olds), ethnically diverse (25% Asian, 8% Black, 58% White, 9% Other) and included 10% (n=86) international students. Response rates were higher among females (11% above the level for UK universities and 14% above the UoL level) and an ethnicity profile intermediate between UoL and UK university undergraduate populations (S3 and S4 Tables). The distribution among year of study was however strongly representative of the UoL population (S5 Table). Two thirds (64%) of students (527/827) reported having had a COVID-19 vaccine at the time of questionnaire completion [74% (390/527) had Pfizer/BioNTech, 23% (121/527) AstraZeneca, 3% (16/527) another vaccine]. A further 194 students (23%) expressed a willingness to become vaccinated, giving a total of 85% who had been or were willing to be vaccinated. Results for 66 students were excluded from further analysis of vaccine hesitancy due to uncertainty about their intention to vaccinate (the selected response was ‘I have not had a vaccination but have been told that I will be offered a vaccination in the near future’). Removing these students from the denominator gave an overall willingness rate of 95% (721/761). There were 40 students (5%) who indicated that they had refused or would refuse a COVID-19 vaccine.

### Univariable analysis of vaccine hesitancy

The results of the univariable analysis for vaccine hesitancy are shown in 40 students (5%) who indicated that they had refused or would refuse a COVID-19 vaccine.

. Ethnicity, course studied, concerns around side-effects (particularly the AstraZeneca vaccine), concerns around spreading COVID-19 to others, place of residence while studying and VAX score were all found to be significantly associated with hesitancy after correcting for multiple testing. There was a weak trend for an association of age with vaccine hesitancy; this could not, however be explored further due to banded collection of age data and the narrow age range of this cohort.

Analysis of the individual VAX scale questions (see S2 Figure) showed that only 29% of hesitant students disagreed that natural exposure to a disease was safer than vaccination compared to 80% of vaccine-willing students. By contrast, 70% of hesitant students, but also 54% of willing students, had concerns about the safety of vaccines (the statement was ‘Although most vaccines appear to be safe, there may be problems that we have not yet discovered’). Approximately half (49%) the hesitant students and 82% of willing students agreed that >95% vaccine coverage was required to prevent the spread of infectious diseases.

A surprising observation was that high proportions of both vaccine-willing (42%, 299/721) and vaccine-hesitant (48%, 19/40) students had experienced a COVID-19 death among relatives or other acquaintances (S3 Figure). This outcome was, however, not associated with differences in hesitancy (Table 1).

**Table 1.**
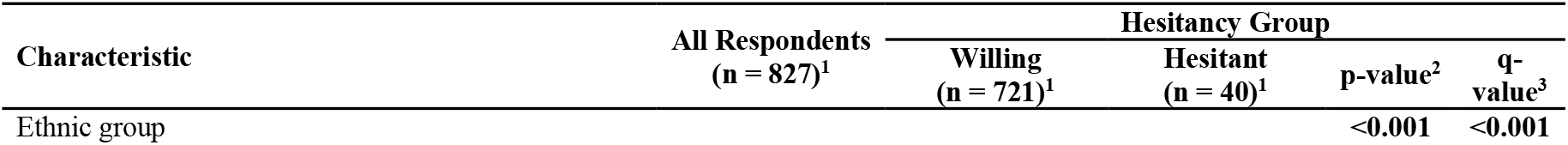
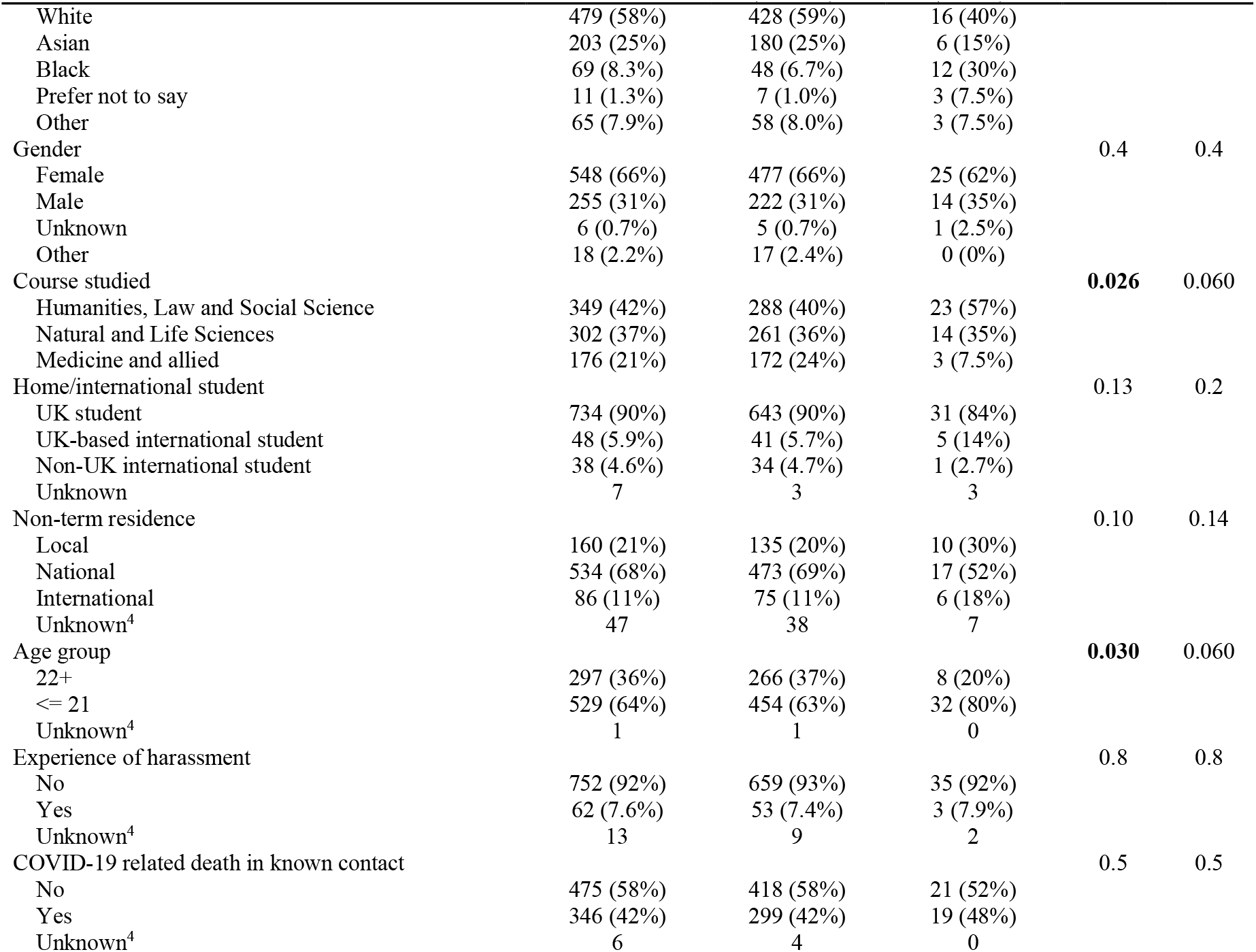
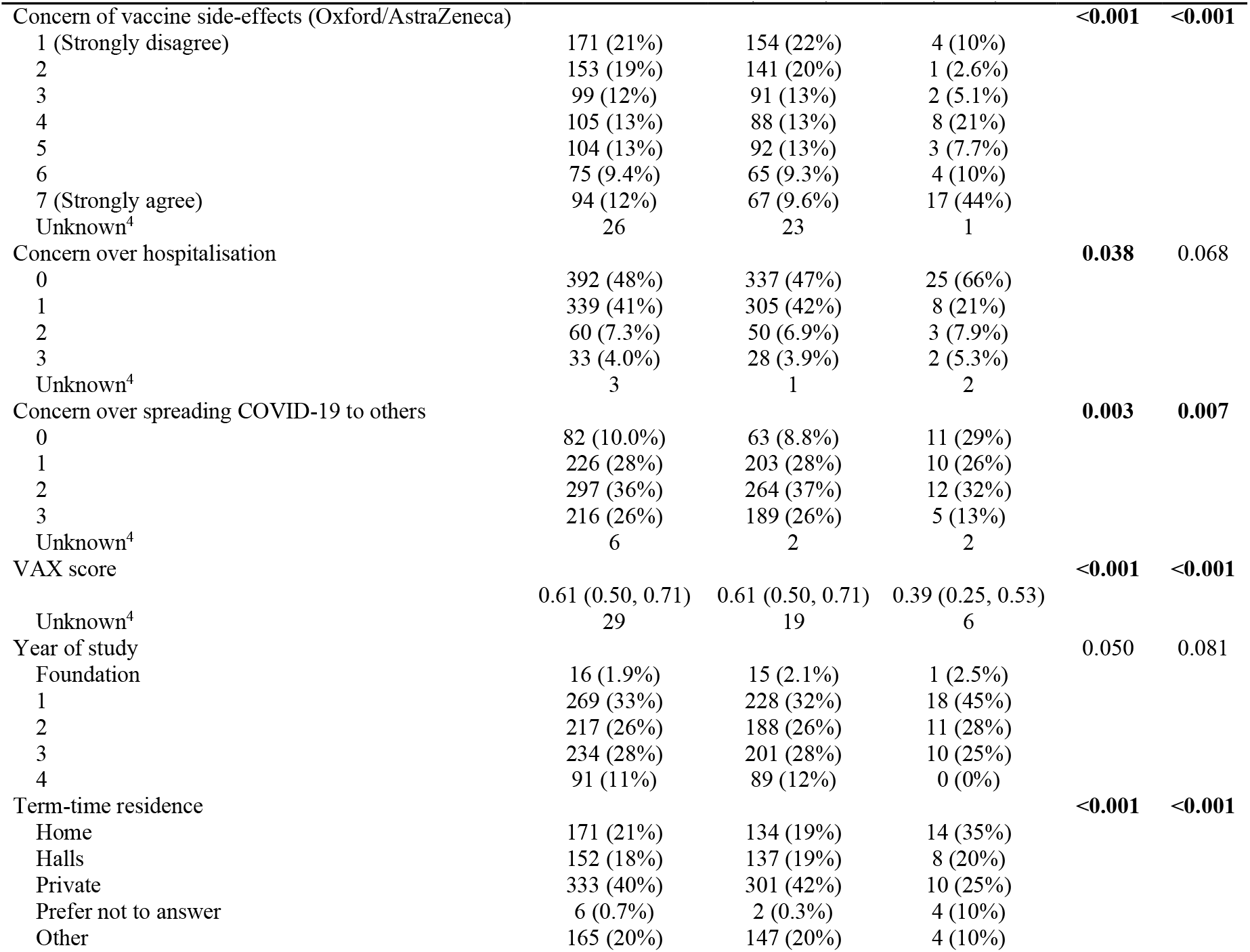
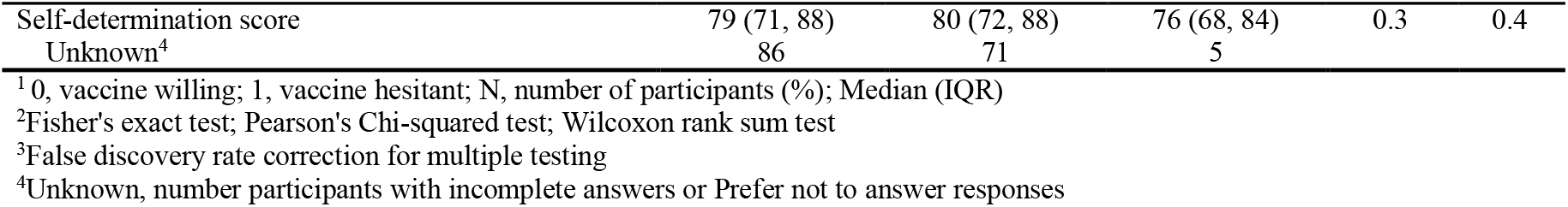
Demographic characteristics of study participants and results of an unweighted univariate analysis of vaccine hesitancy.

### Multivariable analysis of vaccine hesitancy

The multivariable analysis identified associations between vaccine hesitancy and ethnicity, course of study, side-effects and place of term-time residence, as found in the univariate analysis, and additionally with experiences of death among contacts (Table 2). For course studied, studying medicine and allied professions (e.g. midwifery, nursing and physiotherapy) had a significantly lower likelihood of being vaccine hesitant (OR 0.1, 95% CI 0.02-0.5, adjusted P = 0.021) compared to humanities, law and social sciences. For ethnicity, hesitancy among black students had a high odds ratio (OR 7.01, 95% CI 1.81-27.3, adjusted P value = 0.021) as compared to white students (Table 2). Students living in private accommodation (OR 0.13, 95% CI 0.04-0.38, adjusted P=0.004) were less vaccine hesitant than students living at home.

**Table 2.** Multivariate analysis of vaccine hesitancy.

| Characteristic | N | Hesitancy (unweighted) |  |  |  | N | Hesitancy (weighted) |  |  |  |
| --- | --- | --- | --- | --- | --- | --- | --- | --- | --- | --- |
|  |  | OR<sup>1</sup> | 95% CI<sup>1</sup> | p-value | q-value<sup>2</sup> |  | OR<sup>1</sup> | 95% CI<sup>1</sup> | p-value | q-value<sup>2</sup> |
| Gender |  |  |  |  |  |  |  |  |  |  |
| Female | 419 | — | — |  |  | 299 | — | — |  |  |
| Male | 193 | 0.84 | 0.22, 2.89 | 0.8 | >0.9 | 147 | 0.63 | 0.21, 1.85 | 0.4 | 0.6 |
| Unknown | 5 | 0.00 |  | >0.9 | >0.9 |  |  |  |  |  |
| Ethnic group |  |  |  |  |  |  |  |  |  |  |
| White | 377 | — | — |  |  | 271 | — | — |  |  |
| Asian | 145 | 0.23 | 0.04, 1.03 | 0.072 | 0.2 | 97 | 0.24 | 0.04, 1.52 | 0.13 | 0.2 |
| Black | 43 | 6.17 | 1.48, 26.7 | **0.012** | 0.082 | 33 | 7.01 | 1.81, 27.3 | **0.005** | **0.021** |
| Other | 52 | 1.40 | 0.17, 8.22 | 0.7 | >0.9 | 39 | 1.37 | 0.12, 15.4 | 0.8 | >0.9 |
| Age group |  |  |  |  |  |  |  |  |  |  |
| 22+ | 230 | — | — |  |  | 169 | — | — |  |  |
| <= 21 | 387 | 2.92 | 0.78, 12.6 | 0.13 | 0.3 | 276 | 3.25 | 0.73, 14.4 | 0.12 | 0.2 |
| Course studied<sup>3</sup> |  |  |  |  |  |  |  |  |  |  |
| H, Law, Soc | 240 | — | — |  |  | 175 | — | — |  |  |
| Nat/Life Sci | 229 | 1.04 | 0.31, 3.44 | >0.9 | >0.9 | 158 | 0.93 | 0.32, 2.70 | 0.9 | >0.9 |
| Med/Allied | 148 | 0.16 | 0.02, 0.91 | 0.061 | 0.2 | 110 | 0.10 | 0.02, 0.50 | **0.005** | **0.021** |
| Non-term residence |  |  |  |  |  |  |  |  |  |  |
| Local | 120 | — | — |  |  | 92 | — | — |  |  |
| National | 429 | 0.61 | 0.13, 2.79 | 0.5 | 0.8 | 305 | 0.71 | 0.20, 2.46 | 0.6 | 0.7 |
| International | 68 | 1.19 | 0.19, 7.23 | 0.8 | >0.9 | 49 | 1.76 | 0.44, 7.15 | 0.4 | 0.6 |
| Experience of harassment |  |  |  |  |  |  |  |  |  |  |
| No | 570 | — | — |  |  | 413 | — | — |  |  |
| Yes | 47 | 2.42 | 0.34, 13.1 | 0.3 | 0.5 | 31 | 3.61 | 0.51, 25.8 | 0.2 | 0.3 |
| COVID-19 related death in known contact |  |  |  |  |  |  |  |  |  |  |
| No | 351 | — | — |  |  | 267 | — | — |  |  |
| Yes | 266 | 5.05 | 1.65, 17.8 | **0.007** | 0.068 | 197 | 7.49 | 2.06, 27.2 | **0.002** | **0.014** |
| Concern of vaccine side-effects (Oxford/AstraZeneca) |  |  |  |  |  |  |  |  |  |  |
|  | 593 | 1.71 | 1.29, 2.37 | **<0.001** | **0.010** | 610 | 1.72 | 1.23, 2.39 | **0.001** | **0.013** |
| Concern over hospitalisation |  |  |  |  |  |  |  |  |  |  |
|  | 593 | 0.56 | 0.22, 1.25 | 0.2 | 0.3 | 610 | 0.68 | 0.24, 1.89 | 0.5 | 0.6 |

| Characteristic | N | Hesitancy (unweighted) |  |  |  | N | OR<sup>1</sup> | Hesitancy (weighted) |  |  |
| --- | --- | --- | --- | --- | --- | --- | --- | --- | --- | --- |
|  |  | OR<sup>1</sup> | 95% CI<sup>1</sup> | p-value | q-value<sup>2</sup> |  |  | 95% CI<sup>1</sup> | p-value | q-value<sup>2</sup> |
| Concern over spreading COVID-19 to others | 593 | 0.52 | 0.27, 0.96 | **0.041** | 0.14 | 610 | 0.45 | 0.25, 0.81 | **0.008** | **0.024** |
| Year of study | 593 | 0.81 | 0.39, 1.59 | 0.6 | 0.8 | 610 | 0.94 | 0.49, 1.81 | 0.9 | >0.9 |
| Term-time residence |  |  |  |  |  |  |  |  |  |  |
| Home | 124 | — | — |  |  | 93 | — | — |  |  |
| Halls | 119 | 0.23 | 0.04, 1.24 | 0.093 | 0.2 | 96 | 0.20 | 0.04, 1.03 | 0.054 | 0.13 |
| Private | 257 | 0.17 | 0.03, 0.83 | **0.032** | 0.14 | 186 | 0.13 | 0.04, 0.38 | **<0.001** | **0.004** |
| Other | 117 | 0.09 | 0.00, 0.62 | **0.036** | 0.14 | 78 | 0.07 | 0.01, 0.67 | **0.021** | 0.057 |
| Self-determination score | 593 | 1.01 | 0.96, 1.06 | 0.7 | 0.9 | 610 | 1.00 | 0.96, 1.04 | >0.9 | >0.9 |

<sup>1</sup>OR = Odds Ratio, CI = Confidence Interval; <sup>2</sup>False discovery rate correction for multiple testing; 3Abbreviations are:- H, Humanities; Soc, Social Sciences; Nat/Life Sci, Natural and Life Sciences; Med/Allied, Medicine and Allied courses.

Hesitancy was strongly associated with concerns over side-effects from the Astra-Zeneca (Table 2) and Pfizer/BioNTech vaccines (OR 2.1, 95%CI 1.5-3.0, adjusted P<0.001; data not shown). Concerns about side-effects were, however, lower for the Pfizer/BioNTech vaccine (S4 Figure). Surprisingly, the multivariate analysis detected a positive association between experiences of a COVID-19-related death in a family member, friend or other contact with vaccine hesitancy (Table 2). This association remained even when only close contacts (friends; family) were considered (odds ratio 6.4, 95% CI 1.9-21.6, adjusted P = 0.02; data not shown).

### Analysis of VAX scores

Both Asian and Black ethnic groups had significantly lower VAX scores than White ethnicity, indicating a higher level of vaccine hesitancy in the former groups (Figure 1). Similarly, we observed that home students had significantly lower VAX scores than students living in private or other accommodation (Figure 1). The mean VAX score for students living in halls was higher but not significantly different to those living at home, indicating a trend for home students to be more vaccine hesitant than those who lived in other locations during this academic year.

**Figure 1.**
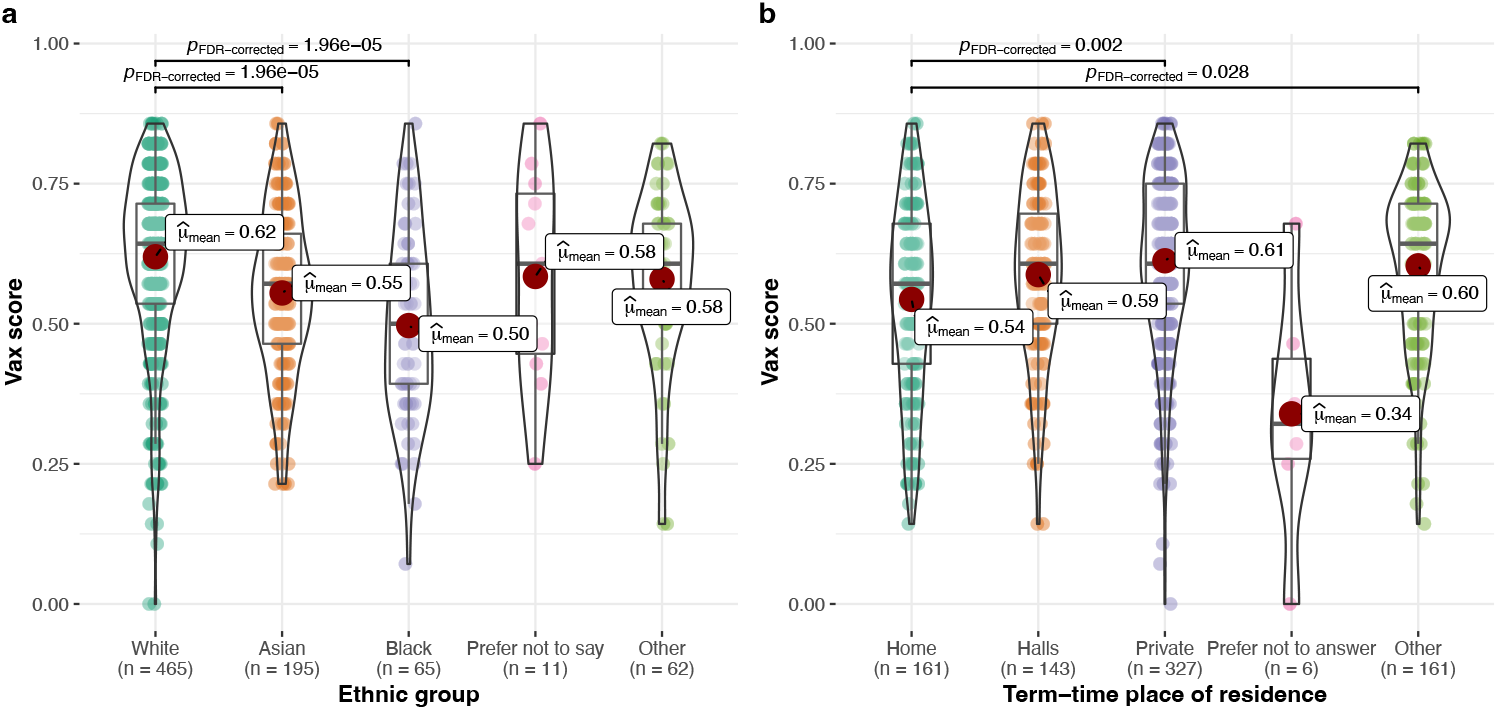
VAX scores for a range of ethnic groups and study residences. The VAX score was determined for each student from responses to four statements about vaccines (statements 17.1, 17.2, 17.3 and 17.5 in Question 17; see S1 Table and S2 Figure). VAX scores range from 0 to 1 representing high to low vaccine hesitancy. The VAX scores were determined for all individuals in four broad ethnic groups (A) or places of residence during the university term (B). Graphs show violin plots with the mean scores indicated by a red circle. Box, IQR; line, IQR + 1.5 times IQR; line within box, median. P values were derived using pairwise tests with FDR correction.

### Knowledge of MMR and MenACWY vaccine status among students

Views on MMR and MenACWY vaccines are shown in Table 3. Very few students (2-4%) self-reported not having had the MMR or MenACWY vaccines, but an additional 8% did not know if they had had their MMR vaccine and 23% did not know if they had had their MenACWY vaccine (Table 3). International students were more likely not to know their vaccination status compared to local students (Table 3). Additionally, 15% of UK students did not know that the MMR and MenACWY were available free of charge in the UK and 6% reported not knowing that COVID-19 vaccines were also available free of charge. Again, these proportions were significantly higher among international students. More than half (57% and 61%, respectively) of students favoured on-campus MMR/MenACWY and COVID-19 vaccine provision, respectively. UK-based international students were also highly supportive of this provision with 52-62% selecting a “definitely increase” response for these vaccines (**Error! Reference source not found**.).

**Table 3.**
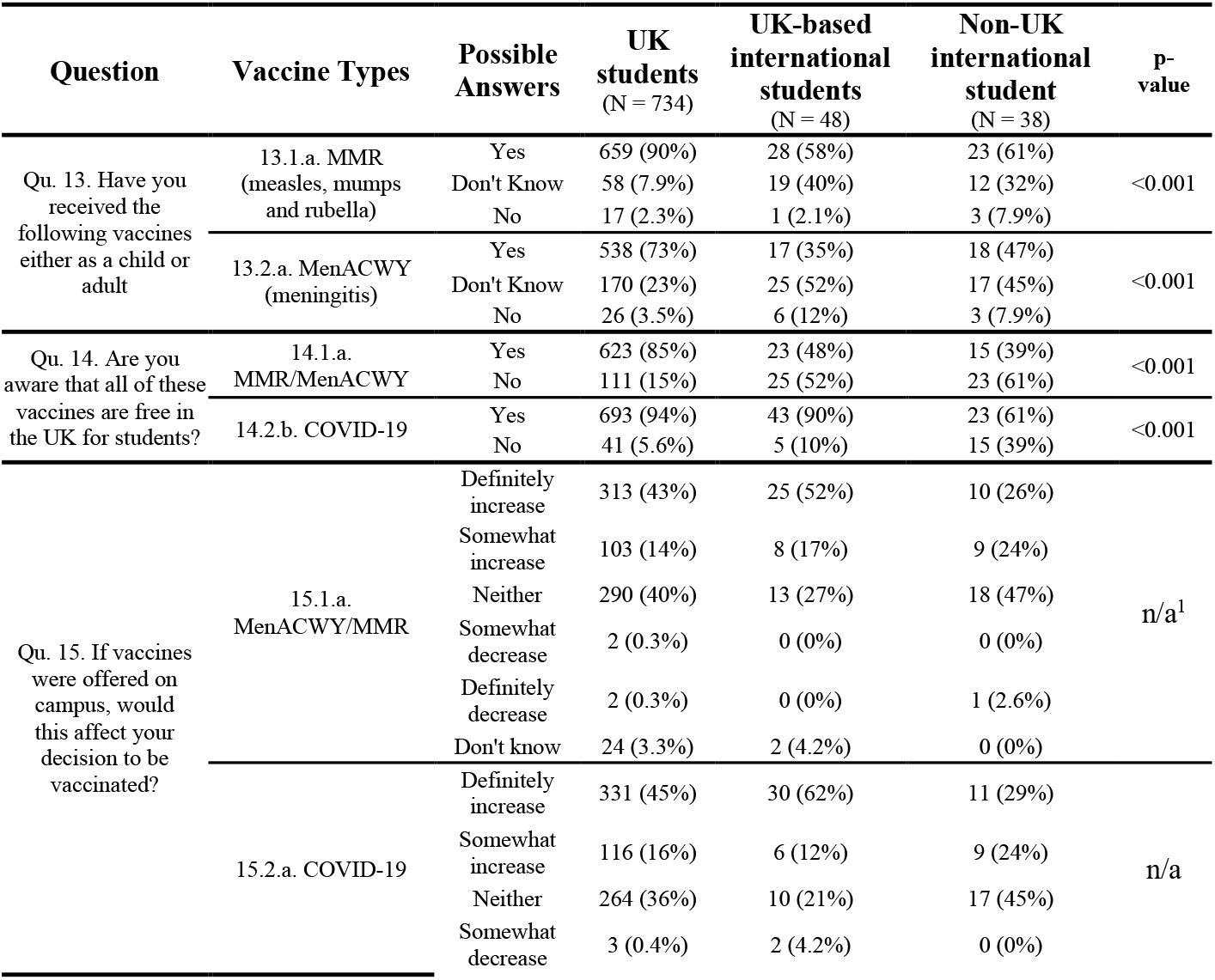
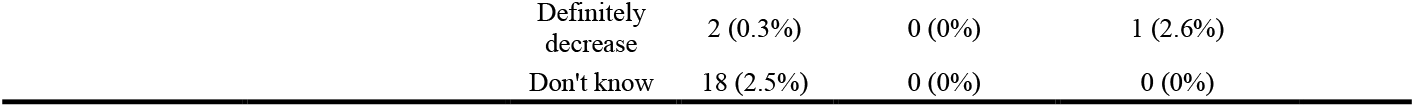
Comparative knowledge and attitudes to on campus delivery of COVID-19, MMR and MenACWY vaccines

### Univariable and multivariable analyses of attitudes to on-campus vaccinations

The univariable analysis of on-campus MMR/MenACWY vaccine programmes identified a significant association with MMR vaccine status indicating that those who responded with either a ‘Yes’ or ‘Don’t Know’ response for their vaccine status were in favour of these programmes (S7 Table). However, these responses were not significant in the multivariate analysis after correction for multiple testing (S8 Table). The univariable analysis of on-campus COVID-19 vaccine provision found significant associations with COVID-19 vaccine hesitancy and term-time residence (S7 Table). In the multivariable analysis, vaccine hesitancy was negatively correlated with on-campus provision (S8 Table). Multivariable logistic regression of term-time residence indicated that students studying in halls (OR 3.5 95% CI 1.6-7.6, adjusted P = 0.021) or private accommodation (OR 2.6, 95% CI 1.3-4.99, adjusted P = 0.03) were in favour of this provision.

## Discussion

University students are a critical group for illness and spread of infectious diseases and hence are an important target for vaccination programmes. Our survey of University of Leicester students was unique in that we evaluated both attitudes to and mechanisms for uptake of the three major vaccines targeted to this population group in the UK. Our study indicates that ethnicity, concerns over side-effects and place of residence are key determinants of COVID-19 vaccine hesitancy. We also found high levels of uncertainty among students about their MenACWY and MMR vaccine status. As an approach to facilitating vaccine uptake, students were asked about on-campus provision of vaccines and reported to favour this approach. Based on our findings we elaborate key recommendations for improved vaccine delivery to this population sector.

Our observed high uptake (64%) of COVID-19 vaccines was surprising given that this age group only became eligible for COVID-19 vaccines during the course of our data collection. These uptake levels were significantly higher than the wider young adult population at that time (P<0.0001 as compared to Leicester 18-24 year olds; S1 Figure), suggesting that these students were more proactive about accessing COVID-19 vaccines than their peers. This high uptake may also have been partially attributable to high participation in the study by pro-vaccine students and/or to surge vaccinations in the Leicester COVID-19 hotspot, including a pop-up vaccination centre in De Montfort Hall, adjacent to the UoL campus, that took place just prior to initiation of the survey. We also observed a high willingness for uptake of COVID-19 vaccines (95%) among the University of Leicester survey respondents. These findings may reflect the effectiveness of both the delivery of vaccines to students and the information campaign on the benefits of these vaccines. The subsequent evidence of uptake rates of >90% in an ONS study of UK university students [32] indicates that, despite the caveats (see below), our study was a reasonable predictor of student attitudes to vaccines. Intriguingly, we also found that 93% (37/40) of the COVID-19 vaccine hesitant individuals reported having had at least one of the MMR and MenACWY vaccines with 50% (20/40) having had both; this would suggest that these individuals are specifically concerned about the COVID-19 vaccines or that vaccine hesitancy has developed during their transition to adulthood.

### Determinants of vaccine hesitancy among university students

A key concern for vaccination programmes has been to identify groups of individuals with lower levels of vaccine uptake. Our univariable analysis indicated that ethnicity was strongly linked to vaccine hesitancy, with the multivariable analysis further linking this hesitancy to the black ethnic group within the wider university student population. As these findings were based on small numbers of vaccine-hesitant individuals, we examined the VAX scores for all individuals of each ethnic group and found that both the Asian and Black ethnic groups had significantly lower average scores than the White ethnic group indicating a general trend towards hesitancy among the minority ethnic groups (Figure 1). Other studies have also found evidence of vaccine hesitancy associated with ethnicity [33-36] and specifically with students of black ethnicity [36]. Hesitancy in these groups has been linked to discrimination, mistrust of healthcare organisations, misinformation, lower perceived vaccine efficacy/safety. A substantial proportion of vaccine hesitant individuals (37.5%; 15/40) in our study agreed with a statement that COVID-19 vaccines had not been thoroughly tested in different ethnic groups (S9 Table).

Our study also identified some evidence that students registered in medical courses were significantly less likely to be vaccine hesitant than students registered for other courses. Other studies have also observed higher uptake of vaccines among medical students [8]. Medical students may be more knowledgeable about vaccines and interested in preventing illnesses, as inculcated in the duty of care for healthcare professionals, and hence are likely to be more in favour of vaccination programmes than other groups.

A novel finding was of an association between vaccine hesitancy and students who lived at home. This association may be part of a general trend as the average VAX scores for all students living at home was significantly lower than those living in private accommodation or other, mainly mixed, accommodation types (Figure 1). Despite lockdowns and extensive online teaching, the home student group was small as only 7% of national and 19% of international students reported remaining at home for the 2019/2020 academic year. Higher vaccine hesitancy in students living at home may be explained by a consideration of the concerns around spreading of COVID-19. While their risk of hospitalization (and hence personal safety) was not a significant predictor of vaccine hesitancy in our multivariate regression (P = 0.6), we found that there were lower levels of vaccine hesitancy among students concerned about spreading COVID-19 (OR 0.5, 95%CI 0.3-0.81, P=0.024). Students living at home were less concerned about spreading COVID-19 than those living away from home (50% and 62%, respectively). We postulate that this attitude may be driven by a lower perceived risk of the potential for spreading the disease due to reduced day-to-day social interactions compared to students living in halls or private accommodation.

A concern raised by our study is of an association between potential side effects of COVID-19 vaccines and vaccine hesitancy. Our study was performed a few months after concerns about side-effects of the AstraZeneca vaccine became apparent, leading to recommendations for young people to receive the Pfizer/BioNTech vaccine instead. Concerns over side-effects were associated with vaccine hesitancy in both our univariate and multivariate models, indicating that this important factor could reduce vaccine uptake. Unfortunately, we did not query which side-effects in particular the students were concerned about and hence it is possible that common but mild side-effects may be enough to dissuade ambivalent students from receiving vaccinations. However, we also note that many students still obtained COVID-19 vaccines despite these concerns suggesting that other factors may over-ride the perceived risks of side-effects. It will be interesting to observe whether this effect has dissipated over time with more information on the low occurrence of severe reactions and improved strategies for preventing occurrences.

### MMR and MenACWY vaccine status

An important strategy for increasing MMR and MenACWY vaccine uptake among young adults is making them aware of their vaccination status [37]. This is now demonstrably possible via digital applications such as the NHSapp and EU Digital COVID certificate. Our survey found high levels of uncertainty among students about their MMR (11% didn’t know) and MenACWY (26% didn’t know) vaccine status and 3-4% who reported no uptake (**Error! Reference source not found**.). In a 2019/2020 questionnaire performed just prior to the pandemic, 16% and 54% of University of Leicester students reported not knowing their MMR and MenACWY vaccine status, respectively [22]. That study also found that 37% of students were unaware that these vaccines were free and accessible through GP practices.

Other estimates of vaccine uptake in England have indicated that 86% of eligible individuals have had both doses of the MMR vaccine (in July 2019) and the single-dose of the MenACWY vaccine (in 2017/2018) with the MMR vaccination level being below the >95% coverage recommend by the World Health Organisation for preventing measles and mumps outbreaks [19, 21, 38]. It is likely that many of the students who are uncertain about their vaccine status have not had these vaccines. A particularly concerning feature of our study was that a higher proportion of international students had not had the MMR and MenACWY or did not know their status for these vaccines; these students will be at a higher potential risk of contracting and/or spreading the diseases targeted by these vaccines. In England, Public Health England recommends that anyone who is uncertain about their vaccine status or has missed a vaccine dose should be offered these vaccines [39]. Most students will be unaware of this recommendation, indeed 20% of UoL students did not know that these vaccines are free (**Error! Reference source not found**.). Addressing these issues is important for maintaining both direct protection and herd protection across the population for measles, mumps and four of the five major serogroups responsible for invasive meningococcal disease in the UK.

### On campus provision of vaccines

A high proportion of students were in favour of on campus provision of MenACWY, MMR and COVID-19 vaccines. These proportions were even higher among international students who were based in the UK, which may reflect difficulties in understanding how to access the UK medical system. The statistically significant evidence of support for provision of on-campus COVID-19 vaccinations by students who are not studying at home is an indication that students value easy access to vaccinations. The success of COVID-19 vaccine pop-ups and the University of Nottingham’s programme for delivery of the MenACWY vaccine in the pre-pandemic era indicate that students value and will access these services [9].

### Recommendations

Harnessing new approaches developed during the UK COVID-19 vaccine roll-out is a potential positive legacy of the pandemic to build-back-better for future generations. Empowering digitally-aware young people to take responsibility for their own health and to engage in community health policies is an achievable, cost-efficient outcome with far-reaching personal and population benefits. A key recommendation is for provision of vaccine status information for all vaccines on digital platforms, such as the NHSapp in the UK. Promoting digital platforms with vaccine status and information about accessing any missed vaccinations should help increase vaccine coverage particular among individuals who are uncertain about their vaccine status. Benefits could include protection of more individuals and improved population protection through reduced transmission. Specific delivery of vaccine information and simple access to vaccines by international students should be a gold-standard for the university sector as the financial gains accrued from these students must be allied to high welfare provision. On-campus vaccination programmes should be widely adopted for all relevant vaccines as the preferred strategy of getting vaccinated prior to attending university has never been optimal.

### Strengths and limitations

A strength of this study is that it is the first to simultaneously evaluate uptake, knowledge and attitudes to COVID-19, MMR and MenACWY vaccines among university students. A further strength is the high number and ethnic diversity of the participant population. The use of multivariable regression was a strength that allowed for adjustment for confounders and for identification of significant associations between variables and vaccination parameters with the potential to inform vaccination policies.

Findings from this study may, however, be affected by the inherent limitations of cross-sectional studies. The response rate of 8% indicates that there may have been a response bias due to demographics. The strengths and limitations arising from a range of demographics were considered above but it is possible that biases from other unaccounted demographics (e.g. socioeconomic status) may confound generalisability of our data to the wider UK student population. A significant potential limitation of our study, and inherent in many studies of vaccination, is enhanced participation by individuals with pro-vaccine attitudes and reduced participation by vaccine hesitant individuals. Our fully anonymised survey system and inducements to participate was designed to minimise this bias. Our level of vaccine hesitancy as determined by vaccine uptake is similar to other studies. However, this strict determination of vaccine hesitancy may have missed the full range of hesitancy and excluded students who obtained the vaccine despite having a degree of vaccine hesitancy.

## Conclusions

The findings from this study indicate that there may be differences in uptake and access to the COVID-19, MenACWY and MMR vaccines among university students. Students of Black ethnicity and those residing at home were less likely to be vaccinated with COVID-19 vaccines. Further research on the reasons for hesitancy may be required in order to delivery more effective, ‘tailored’ vaccine information and to develop methods for enhancing trust and acceptance of vaccines in these groups. High levels of uncertainty about personal vaccine status and availability of the MMR and MenACWY vaccines were observed and are likely to impact on vaccine uptake. On campus vaccination delivery was found to be widely favoured particularly by on-campus and international students. These knowledge gaps and delivery approaches should be considered in future student-focussed vaccination campaigns and explored through additional research. Our findings indicate that adopting ‘best-practices’ of easy access and digital vaccine information within the university-sector may breakdown barriers and future-proof uptake of all required vaccines among students.

## Data Availability

All data produced are available online at https://reshare.ukdataservice.ac.uk/855372/

## Acknowledgements

The authors would like to thank Dan Smith and Tamara Vivian for help with setting up and circulation of the questionnaire to University of Leicester students. The authors would also like to thank all the students who participated in this questionnaire.

## Authors Contributions

The study was conceived and designed by CDB, LG and MP. Data collection was overseen by CDB. Data collation and analysis was performed by AW and CDB. All authors were equally involved in data interpretation and writing of the manuscript.

## Supporting information

**S1 Table. UniCoVac Questionnaire**.

**S2 Table. Weighting values for the weighted multivariate analysis**.

**S3 Table. Comparison of ethnicity demographics of UniCoVac questionnaire participants to University of Leicester and UK universities**

**S4 Table. Comparison of gender demographics of UniCoVac questionnaire participants to University of Leicester and UK universities**

**S5 Table. Comparison of demographics for Year groups between UniCoVac questionnaire participants and University of Leicester undergraduate population**

**S6 Table. Classification of questionnaire respondents for vaccine hesitancy (dichotomous)**^**1**^.

**S7 Table. Univariable analysis of attitudes to on campus delivery of MMR and MenACWY vaccines**

**S8 Table. Multivariate analysis of attitudes to on campus delivery of MMR and MenACWY vaccines**

**S9 Table. Reasons for hesitancy of vaccine hesitant individuals**.

**S1 Figure. Comparison of the vaccination levels of UniCoVac participants to comparator populations**. A total of 527 participants (63.7%) indicated that they had had ‘at least one dose of a COVID-19 vaccine’ in response to question 10 of the UniCoVac questionnaire. This data was analysed relative to the time of submission of the questionnaire to determine whether the proportion of immunised individuals increased during the study and in comparisons to local and countrywide rates for an age-matched population (note that 94% of UniCoVac participants were in the 18-25 age bracket). We note that surge vaccinations were performed in Leicester between 25/05/2021 and 6/06/2021 with 21,500 vaccinations being administered in multiple locations including De Montfort Hall which is adjacent to the University of Leicester campus. Graph **a** shows the accumulated submissions (solid line) for the three weeks that the survey was open and the accumulated number of submissions from vaccinated individuals (dashed line). The submission spikes on the 1^st^, 10^th^ and 16^th^ June equate to days when the questionnaire was sent out to students (indicating that most students answer the survey within a few hours of receipt of the email). There is no substantial difference during the collection period indicating that any effect of the surge vaccinations on uptake occurred prior to but not during the study period. Graph **b** shows the % of vaccinated individuals for the UniCoVac participants (solid black line) and 18-24 year olds in Leicester (dashed blue line), East Midlands (dashed red line) and England (dashed green line). A statistical comparison was performed relative to the Leicester cohort as this cohort has the highest uptake and because a high proportion of University of Leicester students would have been able to take advantage of surge vaccinations in Leicester.

**S2 Figure. Comparison of vaccine confidence scores for vaccine hesitant and non-hesitant participants**. Participants were asked to rank five statements on attitudes to vaccines (the full text of survey question 17 is provided in supplementary information). Ranking was from 1, strongly disagree, to 7, strongly agree. The range of responses for each statement is presented for the hesitant and non-hesitant groups as separate bar graphs with percentages on the left and right indicating disagree (1-3) and agree (5-7) responses, respectively. Note that responses 17.1, 17.2, 17.3 and 17.5 were combined to generate a VAX score.

**S3 Figure. Experiences of deaths among friends, family or others**. Question 21 of the survey asked students: “Do you personally know anyone who has died from COVID-19?” Students were allowed to make multiple selections, but in most cases selected one category. Horizontal columns, selection frequency for each individual option; vertical bars, selection frequency for combinations of options.

**S4 Figure. Concerns about the side-effects of COVID-19 vaccines**. Question 12 asked participants to indicate the extent to which they agreed with a statement on side-effects (i.e. “I am concerned about the side-effects of COVID-19 vaccines?”) for the Oxford AstraZeneca, Pfizer-BioNTech and all COVID-19 vaccines. The extent of agreement was indicated by selecting numbers from 1 (Strongly disagree) to 7 (strongly agree) or Don’t know. Percentages for each score are shown in a bar graph with combined percentage values for scores of 1-3, 4 and 5-7 being shown on the left, middle and right, respectively.

