## Supplementary Tables and Figures for "Cross-Sectional Study of University students’ attitudes to ‘on campus’ delivery of COVID-19 vaccines and future-proofing MenACWY and MMR vaccination rates by adopting COVID-19 vaccine roll-out strategies": Webb_Bayliss_PLOS_ONE-Supplementary_Information-13122021 copy.docx

S1 Table. UniCoVac Questionnaire.

| **Question** | **Answers and Sub-Questions** |
| --- | --- |
| Qu. 2. What is your age group | 16-17, 18-21, 21-25, >25, Prefer not answer |
| Qu. 3. What is your current year of study | Foundation, First, Second, Third, Other, Prefer not to answer |
| Qu. 4. What is the name of your degree course | Open text box |
| Qu. 5. Are you an international student | Yes, No, Prefer not answer. 5a. If Yes please state your country of permanent residence (open text box); 5.b. Are you currently living in the UK (Yes, No, Prefer not to answer) 5.b.i. If Yes please state when you arrived in the UK (Before 2020, January-May 2020, July-December 2020, January-May 2021) |
| Qu.6, What is your gender identity? | Male, Female, Non-binary, Another gender identity, Prefer not to answer |
| Qu. 7. What is your ethnic group? | Asian/Asian British – Indian, Asian/Asian British – Pakistani, Asian/Asian British – Bangladeshi, Asian/Asian British – Chinese, Asian/Asian British – Any other Asian background, Black/African/Caribbean/ Black British – African, Black/African/Caribbean/ Black British – Caribbean, Black/African/Caribbean/ Black, British – Any other Black/African/Caribbean background, Mixed/multiple ethnic groups - White and Black Caribbean, Mixed/multiple ethnic groups - White and Black African, Mixed/multiple ethnic groups - White and Asian, Mixed/multiple ethnic groups - Any other mixed/multiple ethnic background, White – British, White - Irish White - Gypsy or Irish Travellers, White - Any other white background, Other ethnic group – Arab, Other ethnic group – Any other ethnic background, Prefer not to answer |
| Qu. 8. Where have you lived during this academic year (tick all that apply) | Oadby student village, Nixon court, Opal court, Brookland Road, Private student housing, Commute from home, Stayed at home, Other, Prefer not to answer |
| Qu. 9. If you live in the UK, where do you live outside of term time | [please indicate first part of the post-code, eg NG8] |
| Qu. 10. Have you had, or are you going to have, a vaccination against COVID-19? | I have already had at least one COVID-19 vaccination, I have not had a vaccination but have been told that I will be offered a vaccination in the near future, I have been offered a vaccination but have decided not to have the vaccine, I have not yet been offered a vaccination but intend to have the vaccine when offered, I have not yet been offered a vaccination but have decided not to have a vaccine when offered, Other  If selected Other, please specify: (Open text box))  For answers 3 or 5, there was a sub-question:- Qu. 10, b. What are your reason(s) for not having the COVID-19 vaccination? Please select all that apply (for individuals answering b. or d. to Qu. 10). i. I have allergies, needle-phobia, am immuno-compromised, or have other clinical reasons not to be vaccinated; ii. I am not convinced that COVID-19 vaccines will be effective; iii. Vaccines may not have been tested thoroughly in all ethnic groups; iv. I have had COVID-19 and therefore do not feel I need the vaccine; v. I do not feel that I personally am at risk from COVID-19; vi. I do not believe in vaccinations in general; vii. It is inconvenient to get a COVID-19 vaccine; viii. None of the above; viii. Prefer not to answer; ix. Other (If you selected Other, please specify (Open text box))  For answer 1, there was a sub-question: 10.c. Which COVID-19 vaccine did you receive? i. Pfizer-Biontech; ii. Oxford-AstraZeneca; iii. Moderna; iv. Johnson & Johnson; v. Sputnik V; vi. Sinopharm; vii. Sinovac; viii. Unsure; ix. Prefer not to answer; x. Other (If you selected Other, please specify (Open text box)) |
| Qu. 11. How much do you know about COVID-19 vaccines? | indicate on a scale of 1 (low) to 7 (high) your level of knowledge of each of these vaccines:- Pfizer-Biontech; Oxford-AstraZeneca; Moderna; Johnson & Johnson; Sputnik V; Sinopharm; Sinovac |
| Qu. 12. I am concerned about the side-effects of COVID-19 vaccines? | indicate the extent to which you agree or disagree by selecting a number from 1 (Strongly disagree) to 7 (strongly agree) or Don’t know: - Oxford-AstraZeneca, Pfizer-Biontech; All COVID-19 vaccines |
| Qu. 13. Have you received the following vaccines either as a child or adult | a. MMR (measles, mumps and rubella) YES/NO/DON’T KNOW b. MenACWY (meningitis) YES/NO/DON’T KNOW |
| Qu. 14. Are you aware that all of these vaccines are free in the UK for students? | a. MenACWY/MMR, YES/NO; b, COVID-19 vaccines, YES/NO |
| Qu. 15. If vaccines were offered on campus, would this affect your decision to be vaccinated? | a. MenACWY/MMR, Definitely increase /Somewhat increase /Neither/ Somewhat decrease/Definitely decrease/Don’t know; b. COVID-19, Definitely increase /Somewhat increase /Neither /Somewhat decrease/ Definitely decrease/Don’t know |
| Qu. 16. Which location would be the best for accessing vaccines on campus? | Please select up to three with a ranking of 1 (first) to 3 (third); Registration, Fresher’s Fair, Halls of Residence, Student Union. Other main campus building (e.g. Adrian building), Victoria Park Health Centre, Other (If you selected Other, please specify, (open text box)) |
| Qu. 17. How much do you agree with the following statements about vaccinations in general? | indicate the extent to which you agree or disagree by selecting a number from 1 (Strongly disagree) to 7 (strongly agree) or indicate ‘Prefer not to answer’ :-  I can rely on vaccines to stop serious infectious diseases; Although most vaccines appear to be safe, there may be problems that we have not yet discovered; Authorities promote vaccination for financial gain, not for people's health; A high level of vaccine coverage (>95%) is required to prevent spread of infectious diseases; Being exposed to diseases naturally is safer for the immune system than being exposed through vaccination |
| Qu. 18. Do you think that you currently have or have had COVID-19? | No; Unsure; Yes, my own suspicions; Yes, suspected by a doctor but not tested; Yes, confirmed by a positive test; Prefer not to answer. If answer 5 was selected then the following question appeared:- 18.a. Which type of test were/are you positive (PCR, Lateral flow, Both, Unsure, Other). 18a.i. If you selected Other, please specify: (open text box) |
| Qu. 19. How concerned are you that you will get COVID-19 and require hospitalisation? | Not at all concerned; A little concerned; Quite concerned; Very concerned; Prefer not to answer |
| Qu. 20. How concerned are you that you might unknowingly spread COVID-19 to others? | Not at all concerned; A little concerned; Quite concerned; Very concerned; Prefer not to answer |
| Qu. 21. Do you personally know anyone who has died from COVID-19? | (Please select all that apply)  Yes, family member(s); Yes, friend(s); Yes, someone else; No; Prefer not to answer |
| Qu. 22. Where do you get information about COVID-19? (Please select all that apply) | Other students; Friends, family, neighbours; Television, radio, newspapers or magazines; University email or poster; Government or NHS posters, adverts, leaflets or website; Twitter; Tiktok; Other social media (e.g. Facebook, Instagram); World Health Organisation website; Other websites; Scientific journals; Not applicable; Prefer not to answer |
| Qu. 23. Have you experienced any harassment as a result of COVID-19? **This could take the form of verbal abuse, intimidatory or threatening behaviour, receiving hostile messages online, unwanted sexual contact or physical attacks.** | Yes, No, Prefer not to answer If a Yes answer was provided the following questions appeared:- 24. What kind of harassment have you experienced as a result of COVID-19? (Verbally abused; Threatened or intimidated in person; Threatened or intimidated online; Deliberate damage to property; Physically attacked; Sexually harassed or assaulted); 25. What do you think was the main reason for the harassment you have experienced as a result of COVID-19? **Please note that you can be targeted for your perceived association to a group, whether or not you actually identify as a member of it. (**Your age; Your national origins; Your race; Your language or accent; Your religion; An aspect of your physical appearance; Other ( If you selected Other, please specify: (Open text box)); Prefer not to answer |
| Qu. 26. For each of the following statements, indicate the extent to which you agree or disagree by selecting a number from 1 (Strongly disagree) to 7 (strongly agree) or Prefer not to answer. | i. My life is determined by my own actions; ii. I am usually able to protect my personal interests; iii. I can pretty much determine what will happen in my life; iv. To a great extent, my life is controlled by accidental happenings; v. Often there is no chance of protecting my personal interest from bad luck happenings; vi. When I get what I want, it's usually because I'm lucky; vii. People like myself have very little chance of protecting our personal interests where they conflict with those of strong pressure groups; viii. My life is chiefly controlled by powerful others; ix. I feel like what happens in my life is mostly determined by powerful people; x. If someone is meant to have a serious disease, they will get that disease; xi. My health is determined by fate; xii. My health is determined by something greater than myself; xiii. I will stay healthy if I am lucky; xiv. I like having a clear and structured mode of life; xv. I would quickly become impatient and irritated if I could not find a solution to a problem immediately; xvi. Genes are more important than one's own behaviour in determining one's health |
| Qu. 27. How satisfied or dissatisfied are you with the support you received from the University of Leicester during the COVID-19 pandemic | Very satisfied; Moderately satisfied; Neither; Moderately dissatisfied; Very dissatisfied; Prefer not to answer |
| Qu. 28. Is there anything specific that the University of Leicester could do to mitigate the impact on students of any future COVID-19 waves? | Open text box |
| Qu. 29. Would you be willing to be contacted by the project team to  discuss your experiences in more  detail? | Yes, No |

S2 Table. Weighting values for the weighted multivariate analysis.

| **Ethnic_group** | **Gender** | **Weight** |
| --- | --- | --- |
| White | Female | 1.0963362 |
| White | Male | 1.7224416 |
| White | Other | 0.0782102 |
| Asian | Female | 0.4013974 |
| Asian | Male | 0.6306310 |
| Asian | Other | 0.0286348 |
| Black | Female | 0.8557532 |
| Black | Male | 1.3444643 |
| Black | Other | 0.0610475 |
| Prefer not to say | Female | 1.0961934 |
| Prefer not to say | Male | 1.7222171 |
| Other | Female | 0.6639342 |
| Other | Male | 1.0430996 |
| Other | Other | 0.0473636 |

S3 Table. Comparison of ethnicity demographics of UniCoVac questionnaire participants to University of Leicester and UK universities

| **Ethnic group** | **Survey** | **University of Leicester^1^** | **UK universities^1^** |
| --- | --- | --- | --- |
| White | 479 (58%) | 4,798 (41%) | 1,444,450 (74%) |
| Asian | 203 (25%) | 4,055 (34%) | 226,595 (12%) |
| Black | 69 (8%) | 1,552 (13%) | 152,420 (8%) |
| Other | 65 (8%) | 794 (7%) | 118,250 (8%) |
| (Unknown) | 11 (1%) | 540 (5%) | 33,660 (2%) |

^1^Ethnicity distribution for UniCoVac survey sample is distinct from both the University of Leicester and UK Universities (Chi-squared, 3 d.f., p < 0.00001)

S4 Table. Comparison of gender demographics of UniCoVac questionnaire participants to University of Leicester and UK universities

| **Gender** | **Survey** | **University of Leicester^1^** | **UK universities^1^** |
| --- | --- | --- | --- |
| Female | 548 (66%) | 6,109 (52%) | 1,440,815 (57%) |
| Male | 255 (31%) | 5,630 (48%) | 1,087,710 (43%) |
| Other | 18 (2%) | - | 3,865 (0.2%) |
| Unknown | 6 (1%) | - | - |

^1^Gender distribution for survey sample is distinct from both the University of Leicester and UK Universities (Chi-squared, 1 d.f., p < 0.00001), with an over-representation of female students

S5 Table. Comparison of demographics for Year groups between UniCoVac questionnaire participants and University of Leicester undergraduate population

| **Year of study** | **Survey** | **University^1^** |
| --- | --- | --- |
| Foundation | 16 (2%) | 550 (5%) |
| First | 269 (33%) | 3760 (33%) |
| Second | 217 (26%) | 3298 (29%) |
| Third | 234 (28%) | 2951 (26%) |
| Other | 91 (11%) | 741 (7%) |

^1^Chi-squared test: p = 0.22

S6 Table. Classification of questionnaire respondents for vaccine hesitancy (dichotomous)^1^.

| **Classification and Response to Survey Qu. 10** | **UK/International Student Status** | | | | **Total** |
| --- | --- | --- | --- | --- | --- |
|  | UK student | UK-based international student | Non-UK international student | No answer |  |
| **Not hesitant**  I have already had at least one COVID-19 vaccination | 475 (65%) | 26 (54%) | 25 (66%) | 1 (14%) | 527 (64%) |
| **Hesitant**  I have been offered a vaccination but have decided not to have the vaccine | 18 (2.5%) | 3 (6.2%) | 0 (0%) | 1 (14%) | 22 (2.7%) |
| **Unknown**  I have not had a vaccination but have been told that I will be offered a vaccination in the near future | 49 (6.7%) | 2 (4.2%) | 2 (5.3%) | 1 (14%) | 54 (6.5%) |
| **Hesitant**  I have not yet been offered a vaccination but have decided not to have a vaccine when offered | 13 (1.8%) | 2 (4.2%) | 1 (2.6%) | 2 (29%) | 18 (2.2%) |
| **Not hesitant**  I have not yet been offered a vaccination but intend to have the vaccine when offered | 168 (23%) | 15 (31%) | 9 (24%) | 2 (29%) | 194 (23%) |
| **Unknown**  Other | 11 (1.5%) | 0 (0%) | 1 (2.6%) | 0 (0%) | 12 (1.5%) |
| Total | 734 (100%) | 48 (100%) | 38 (100%) | 7 (100%) | 827 (100%) |

^1^Students were classed as vaccine hesitant / not hesitant or unknown according to their answers to the question “10. Have you had, or are you going to have, a vaccination against COVID-19 (Please select one)?”.

S7 Table. Univariable analysis of attitudes to on campus delivery of MMR and MenACWY vaccines

| Characteristic | **On-campus COVID19 vaccinations** | | | | **On-campus MMR vaccinations** | | | |
| --- | --- | --- | --- | --- | --- | --- | --- | --- |
|  | Ambivalent  (n = 320)^1^ | In Favour  (n = 507)^1^ | p-value^2^ | q-value^3^ | Ambivalent  (n = 355)^1^ | In Favour  (n = 472)^1^ | p-value^2^ | q-value^3^ |
| COVID19 vaccine hesitancy |  |  | **<0.001** | **<0.001** |  |  | **<0.001** | **0.002** |
| 0 | 260 (36%) | 461 (64%) |  |  | 292 (40%) | 429 (60%) |  |  |
| 1 | 30 (75%) | 10 (25%) |  |  | 28 (70%) | 12 (30%) |  |  |
| Unknown | 30 | 36 |  |  | 35 | 31 |  |  |
| Ethnic group |  |  | >0.9 | >0.9 |  |  | 0.7 | 0.7 |
| White | 185 (39%) | 294 (61%) |  |  | 196 (41%) | 283 (59%) |  |  |
| Asian | 78 (38%) | 125 (62%) |  |  | 90 (44%) | 113 (56%) |  |  |
| Black | 28 (41%) | 41 (59%) |  |  | 33 (48%) | 36 (52%) |  |  |
| Prefer not to say | 3 (27%) | 8 (73%) |  |  | 5 (45%) | 6 (55%) |  |  |
| Other | 26 (40%) | 39 (60%) |  |  | 31 (48%) | 34 (52%) |  |  |
| Gender |  |  | 0.8 | >0.9 |  |  | 0.3 | 0.4 |
| Female | 209 (38%) | 339 (62%) |  |  | 231 (42%) | 317 (58%) |  |  |
| Male | 102 (40%) | 153 (60%) |  |  | 115 (45%) | 140 (55%) |  |  |
| Unknown | 3 (50%) | 3 (50%) |  |  | 4 (67%) | 2 (33%) |  |  |
| Other | 6 (33%) | 12 (67%) |  |  | 5 (28%) | 13 (72%) |  |  |
| Course studied |  |  | 0.5 | 0.7 |  |  | 0.4 | 0.4 |
| Humanities, Law and Social Science | 127 (36%) | 222 (64%) |  |  | 148 (42%) | 201 (58%) |  |  |
| Natural and Life Sciences | 123 (41%) | 179 (59%) |  |  | 138 (46%) | 164 (54%) |  |  |
| Medicine and allied | 70 (40%) | 106 (60%) |  |  | 69 (39%) | 107 (61%) |  |  |
| Home / international student |  |  | 0.080 | 0.2 |  |  | 0.2 | 0.3 |
| UK student | 287 (39%) | 447 (61%) |  |  | 318 (43%) | 416 (57%) |  |  |
| UK-based international student | 12 (25%) | 36 (75%) |  |  | 15 (31%) | 33 (69%) |  |  |
| Non-UK international student | 18 (47%) | 20 (53%) |  |  | 19 (50%) | 19 (50%) |  |  |
| Unknown | 3 | 4 |  |  | 3 | 4 |  |  |
| Non-term residence |  |  | 0.090 | 0.2 |  |  | 0.3 | 0.4 |
| Local | 74 (46%) | 86 (54%) |  |  | 77 (48%) | 83 (52%) |  |  |
| National | 199 (37%) | 335 (63%) |  |  | 224 (42%) | 310 (58%) |  |  |
| International | 30 (35%) | 56 (65%) |  |  | 34 (40%) | 52 (60%) |  |  |
| Unknown | 17 | 30 |  |  | 20 | 27 |  |  |
| Age group |  |  | 0.092 | 0.2 |  |  | 0.4 | 0.4 |
| 22+ | 126 (42%) | 171 (58%) |  |  | 133 (45%) | 164 (55%) |  |  |
| <= 21 | 193 (36%) | 336 (64%) |  |  | 221 (42%) | 308 (58%) |  |  |
| Unknown | 1 | 0 |  |  | 1 | 0 |  |  |
| Experience of harassment |  |  | **0.016** | 0.093 |  |  | **0.023** | 0.088 |
| No | 299 (40%) | 453 (60%) |  |  | 330 (44%) | 422 (56%) |  |  |
| Yes | 15 (24%) | 47 (76%) |  |  | 18 (29%) | 44 (71%) |  |  |
| Unknown | 6 | 7 |  |  | 7 | 6 |  |  |
| COVID-19 related death in known contact |  |  | 0.4 | 0.6 |  |  | 0.4 | 0.4 |
| No | 177 (37%) | 298 (63%) |  |  | 196 (41%) | 279 (59%) |  |  |
| Yes | 139 (40%) | 207 (60%) |  |  | 154 (45%) | 192 (55%) |  |  |
| Unknown | 4 | 2 |  |  | 5 | 1 |  |  |
| Concern of vaccine side-effects (Oxford/AstraZeneca) |  |  | 0.9 | >0.9 |  |  | 0.12 | 0.3 |
| 1 | 71 (42%) | 100 (58%) |  |  | 75 (44%) | 96 (56%) |  |  |
| 2 | 63 (41%) | 90 (59%) |  |  | 64 (42%) | 89 (58%) |  |  |
| 3 | 27 (27%) | 72 (73%) |  |  | 29 (29%) | 70 (71%) |  |  |
| 4 | 38 (36%) | 67 (64%) |  |  | 36 (34%) | 69 (66%) |  |  |
| 5 | 36 (35%) | 68 (65%) |  |  | 46 (44%) | 58 (56%) |  |  |
| 6 | 31 (41%) | 44 (59%) |  |  | 38 (51%) | 37 (49%) |  |  |
| 7 | 44 (47%) | 50 (53%) |  |  | 51 (54%) | 43 (46%) |  |  |
| Unknown | 10 | 16 |  |  | 16 | 10 |  |  |
| Concern over hospitalisation |  |  | 0.7 | 0.8 |  |  | >0.9 | >0.9 |
| 0 | 160 (41%) | 232 (59%) |  |  | 169 (43%) | 223 (57%) |  |  |
| 1 | 125 (37%) | 214 (63%) |  |  | 144 (42%) | 195 (58%) |  |  |
| 2 | 21 (35%) | 39 (65%) |  |  | 26 (43%) | 34 (57%) |  |  |
| 3 | 12 (36%) | 21 (64%) |  |  | 14 (42%) | 19 (58%) |  |  |
| Unknown | 2 | 1 |  |  | 2 | 1 |  |  |
| Concern over spreading COVID-19 to others |  |  | 0.4 | 0.6 |  |  | 0.2 | 0.3 |
| 0 | 35 (43%) | 47 (57%) |  |  | 38 (46%) | 44 (54%) |  |  |
| 1 | 95 (42%) | 131 (58%) |  |  | 109 (48%) | 117 (52%) |  |  |
| 2 | 106 (36%) | 191 (64%) |  |  | 118 (40%) | 179 (60%) |  |  |
| 3 | 81 (38%) | 135 (62%) |  |  | 87 (40%) | 129 (60%) |  |  |
| Unknown | 3 | 3 |  |  | 3 | 3 |  |  |
| VAX score | 0.61 (0.50, 0.71) | 0.61 (0.50, 0.71) | 0.6 | 0.8 | 0.61 (0.50, 0.71) | 0.61 (0.50, 0.71) | **0.039** | 0.12 |
| Unknown | 17 | 12 |  |  | 17 | 12 |  |  |
| Year of study |  |  | 0.4 | 0.6 |  |  | 0.4 | 0.4 |
| 0 | 5 (31%) | 11 (69%) |  |  | 5 (31%) | 11 (69%) |  |  |
| 1 | 111 (41%) | 158 (59%) |  |  | 124 (46%) | 145 (54%) |  |  |
| 2 | 73 (34%) | 144 (66%) |  |  | 83 (38%) | 134 (62%) |  |  |
| 3 | 93 (40%) | 141 (60%) |  |  | 104 (44%) | 130 (56%) |  |  |
| 4 | 38 (42%) | 53 (58%) |  |  | 39 (43%) | 52 (57%) |  |  |
| Term-time residence |  |  | **<0.001** | **0.004** |  |  | **<0.001** | **<0.001** |
| Home | 93 (54%) | 78 (46%) |  |  | 100 (58%) | 71 (42%) |  |  |
| Halls | 48 (32%) | 104 (68%) |  |  | 58 (38%) | 94 (62%) |  |  |
| Private | 118 (35%) | 215 (65%) |  |  | 128 (38%) | 205 (62%) |  |  |
| Prefer not to answer | 6 (100%) | 0 (0%) |  |  | 6 (100%) | 0 (0%) |  |  |
| Other | 55 (33%) | 110 (67%) |  |  | 63 (38%) | 102 (62%) |  |  |
| Self-determination score | 80 (72, 87) | 79 (71, 88) | 0.4 | 0.6 | 80 (71, 88) | 79 (72, 87) | 0.3 | 0.4 |
| Unknown | 31 | 55 |  |  | 44 | 42 |  |  |
| MMR vaccination at childhood^4^ |  |  | **0.007** | 0.064 |  |  | **0.002** | **0.010** |
| Yes | 286 (40%) | 427 (60%) |  |  | 308 (43%) | 405 (57%) |  |  |
| No | 11 (52%) | 10 (48%) |  |  | 16 (76%) | 5 (24%) |  |  |
| Don't Know | 23 (25%) | 70 (75%) |  |  | 31 (33%) | 62 (67%) |  |  |
| MenACWY vaccination at childhood^4^ |  |  | 0.061 | 0.2 |  |  | 0.078 | 0.2 |
| Yes | 230 (40%) | 345 (60%) |  |  | 260 (45%) | 315 (55%) |  |  |
| No | 18 (51%) | 17 (49%) |  |  | 16 (46%) | 19 (54%) |  |  |
| Don't Know | 72 (33%) | 145 (67%) |  |  | 79 (36%) | 138 (64%) |  |  |
| ^1^n (%); Median (IQR) | | | | | | | | |
| ^2^Pearson's Chi-squared test; Fisher's exact test; Wilcoxon rank sum test | | | | | | | | |
| ^3^False discovery rate correction for multiple testing ^4^Response groups: In favour = Definitely or somewhat increase, Ambivalent = neither, definitely or somewhat decrease  ^5^ND, no data | | | | | | | | |

S8 Table. Multivariate analysis of attitudes to on campus delivery of MMR and MenACWY vaccines

|  | **On-campus COVID-19**  (weighted) | | | | | **On-campus MenACWY / MMR**  (weighted) | | | | |
| --- | --- | --- | --- | --- | --- | --- | --- | --- | --- | --- |
| Characteristic | N | OR^1^ | 95% CI^1^ | p-value | q-value^2^ | N | OR^1^ | 95% CI^1^ | p-value | q-value^2^ |
| COVID19 vaccine hesitancy |  |  |  |  |  |  |  |  |  |  |
| 0 | 411 | — | — |  |  | 411 | — | — |  |  |
| 1 | 13 | 0.08 | 0.02, 0.44 | **0.003** | **0.030** | 13 | 0.51 | 0.12, 2.22 | 0.4 | 0.6 |
| Gender |  |  |  |  |  |  |  |  |  |  |
| Female | 280 | — | — |  |  | 280 | — | — |  |  |
| Male | 142 | 0.93 | 0.61, 1.41 | 0.7 | 0.9 | 142 | 0.87 | 0.58, 1.32 | 0.5 | 0.6 |
| Ethnic group |  |  |  |  |  |  |  |  |  |  |
| White | 257 | — | — |  |  | 257 | — | — |  |  |
| Asian | 104 | 1.30 | 0.78, 2.17 | 0.3 | 0.5 | 104 | 1.20 | 0.74, 1.95 | 0.5 | 0.6 |
| Black | 35 | 2.80 | 1.12, 7.03 | **0.028** | 0.11 | 35 | 1.67 | 0.72, 3.86 | 0.2 | 0.4 |
| Other | 35 | 1.01 | 0.51, 2.02 | >0.9 | >0.9 | 35 | 0.79 | 0.40, 1.57 | 0.5 | 0.6 |
| Age group |  |  |  |  |  |  |  |  |  |  |
| 22+ | 173 | — | — |  |  | 173 | — | — |  |  |
| <= 21 | 271 | 1.73 | 1.09, 2.76 | **0.021** | 0.11 | 271 | 1.44 | 0.90, 2.31 | 0.12 | 0.3 |
| Course studied |  |  |  |  |  |  |  |  |  |  |
| Humanities, Law and Social Science | 166 | — | — |  |  | 166 | — | — |  |  |
| Natural and Life Sciences | 163 | 0.64 | 0.40, 1.02 | 0.060 | 0.2 | 163 | 0.66 | 0.41, 1.04 | 0.072 | 0.2 |
| Medicine and allied | 110 | 0.88 | 0.51, 1.53 | 0.7 | 0.9 | 110 | 0.97 | 0.57, 1.65 | >0.9 | >0.9 |
| Non-term residence |  |  |  |  |  |  |  |  |  |  |
| Local | 80 | — | — |  |  | 80 | — | — |  |  |
| National | 304 | 0.91 | 0.49, 1.68 | 0.8 | 0.9 | 304 | 0.95 | 0.52, 1.74 | 0.9 | >0.9 |
| International | 49 | 0.68 | 0.29, 1.60 | 0.4 | 0.6 | 49 | 0.75 | 0.33, 1.71 | 0.5 | 0.6 |
| Experience of harassment |  |  |  |  |  |  |  |  |  |  |
| No | 407 | — | — |  |  | 407 | — | — |  |  |
| Yes | 36 | 2.03 | 0.91, 4.51 | 0.083 | 0.2 | 36 | 2.23 | 0.98, 5.10 | 0.057 | 0.2 |
| COVID-19 related death in known contact |  |  |  |  |  |  |  |  |  |  |
| No | 245 | — | — |  |  | 245 | — | — |  |  |
| Yes | 183 | 0.79 | 0.52, 1.18 | 0.2 | 0.5 | 183 | 0.74 | 0.50, 1.09 | 0.13 | 0.3 |
| Concern of vaccine side-effects (Oxford/AstraZeneca) | 599 | 0.94 | 0.85, 1.05 | 0.3 | 0.5 | 599 | 0.88 | 0.79, 0.98 | **0.024** | 0.14 |
| Concern over hospitalisation | 599 | 1.07 | 0.79, 1.44 | 0.7 | 0.9 | 599 | 0.94 | 0.71, 1.24 | 0.7 | 0.7 |
| Concern over spreading COVID-19 to others | 599 | 1.06 | 0.84, 1.33 | 0.6 | 0.9 | 599 | 1.18 | 0.94, 1.48 | 0.2 | 0.3 |
| VAX score | 599 | 0.38 | 0.08, 1.66 | 0.2 | 0.4 | 599 | 0.36 | 0.08, 1.54 | 0.2 | 0.3 |
| Year of study | 599 | 1.21 | 0.95, 1.52 | 0.12 | 0.3 | 599 | 1.12 | 0.88, 1.41 | 0.4 | 0.6 |
| Term-time residence |  |  |  |  |  |  |  |  |  |  |
| Home | 86 | — | — |  |  | 86 | — | — |  |  |
| Halls | 74 | 3.50 | 1.61, 7.61 | **0.002** | **0.021** | 74 | 2.32 | 1.09, 4.94 | **0.030** | 0.14 |
| Private | 173 | 2.59 | 1.34, 4.99 | **0.005** | **0.030** | 173 | 2.36 | 1.24, 4.46 | **0.009** | 0.11 |
| Prefer not to answer | 1 | 0.00 | 0.00, 0.00 | **<0.001** | **<0.001** | 1 | 0.00 | 0.00, 0.00 | **<0.001** | **<0.001** |
| Other | 79 | 2.15 | 1.08, 4.29 | **0.029** | 0.11 | 79 | 2.10 | 1.06, 4.16 | **0.032** | 0.14 |
| Self-determination score | 599 | 1.00 | 0.98, 1.02 | >0.9 | >0.9 | 599 | 0.99 | 0.98, 1.01 | 0.4 | 0.6 |
| MMR vaccination at childhood |  |  |  |  |  |  |  |  |  |  |
| Yes | 398 | — | — |  |  | 398 | — | — |  |  |
| No | 6 | 0.64 | 0.06, 7.16 | 0.7 | 0.9 | 6 | 0.15 | 0.03, 0.82 | **0.028** | 0.14 |
| Don't Know | 45 | 2.59 | 1.03, 6.55 | **0.044** | 0.14 | 45 | 0.98 | 0.43, 2.24 | >0.9 | >0.9 |
| MenACWR vaccination at childhood |  |  |  |  |  |  |  |  |  |  |
| Yes | 310 | — | — |  |  | 310 | — | — |  |  |
| No | 14 | 1.11 | 0.28, 4.41 | 0.9 | >0.9 | 14 | 3.03 | 0.72, 12.7 | 0.13 | 0.3 |
| Don't Know | 115 | 1.03 | 0.62, 1.72 | >0.9 | >0.9 | 115 | 1.76 | 1.03, 2.98 | **0.037** | 0.14 |
| ^1^OR = Odds Ratio, CI = Confidence Interval | | | | | | | | | | |
| ^2^False discovery rate correction for multiple testing | | | | | | | | | | |

S9 Table. Reasons for hesitancy of vaccine hesitant individuals.

| Response | Number Individuals | Percentage of Individuals |
| --- | --- | --- |
| 1.     I have allergies, needle-phobia, am immuno-compromised, or have other clinical reasons not to be vaccinated | 4 | 10% |
| 2.     I am not convinced that COVID-19 vaccines will be effective | 17 | 42.5% |
| 3.     Vaccines may not have been tested thoroughly in all ethnic groups | 15 | 37.5% |
| 4.     I have had COVID-19 and therefore do not feel I need the vaccine | 4 | 10% |
| 5.     I do not feel that I personally am at risk from COVID-19 | 13 | 32.5% |
| 6.     I do not believe in vaccinations in general | 3 | 7.5% |
| 7.     It is inconvenient to get a COVID-19 vaccine | 8 | 20% |
| 8.     None of the above | 5 | 12.5% |
| 9.     Prefer not to answer | 2 | 5% |
| 10.  Other | 8 | 20% |

^1^Individuals who indicated that they would not take up a COVID-19 vaccine (n=40) were asked to ask a sub-question; in the sub-question they could select multiple responses from 10 statements as their reason for not having a COVID-19 vaccination

**Supplementary Figures**

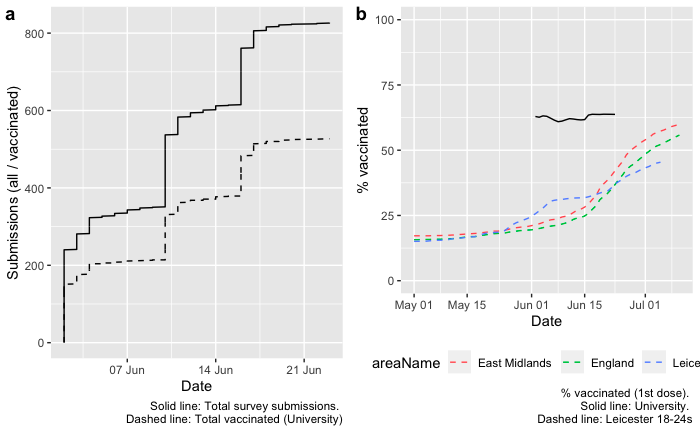

**S1 Figure. Comparison of the vaccination levels of UniCoVac participants to comparator populations.** A total of 527 participants (63.7%) indicated that they had had ‘at least one dose of a COVID-19 vaccine’ in response to question 10 of the UniCoVac questionnaire. This data was analysed relative to the time of submission of the questionnaire to determine whether the proportion of immunised individuals increased during the study and in comparisons to local and countrywide rates for an age-matched population (note that 94% of UniCoVac participants were in the 18-25 age bracket). We note that surge vaccinations were performed in Leicester between 25/05/2021 and 6/06/2021 with 21,500 vaccinations being administered in multiple locations including De Montfort Hall which is adjacent to the University of Leicester campus. Graph **a** shows the accumulated submissions (solid line) for the three weeks that the survey was open and the accumulated number of submissions from vaccinated individuals (dashed line). The submission spikes on the 1^st^, 10^th^ and 16^th^ June equate to days when the questionnaire was sent out to students (indicating that most students answer the survey within a few hours of receipt of the email). There is no substantial difference during the collection period indicating that any effect of the surge vaccinations on uptake occurred prior to but not during the study period. Graph **b** shows the % of vaccinated individuals for the UniCoVac participants (solid black line) and 18-24 year olds in Leicester (dashed blue line), East Midlands (dashed red line) and England (dashed green line). A statistical comparison was performed relative to the Leicester cohort as this cohort has the highest uptake and because a high proportion of University of Leicester students would have been able to take advantage of surge vaccinations in Leicester.

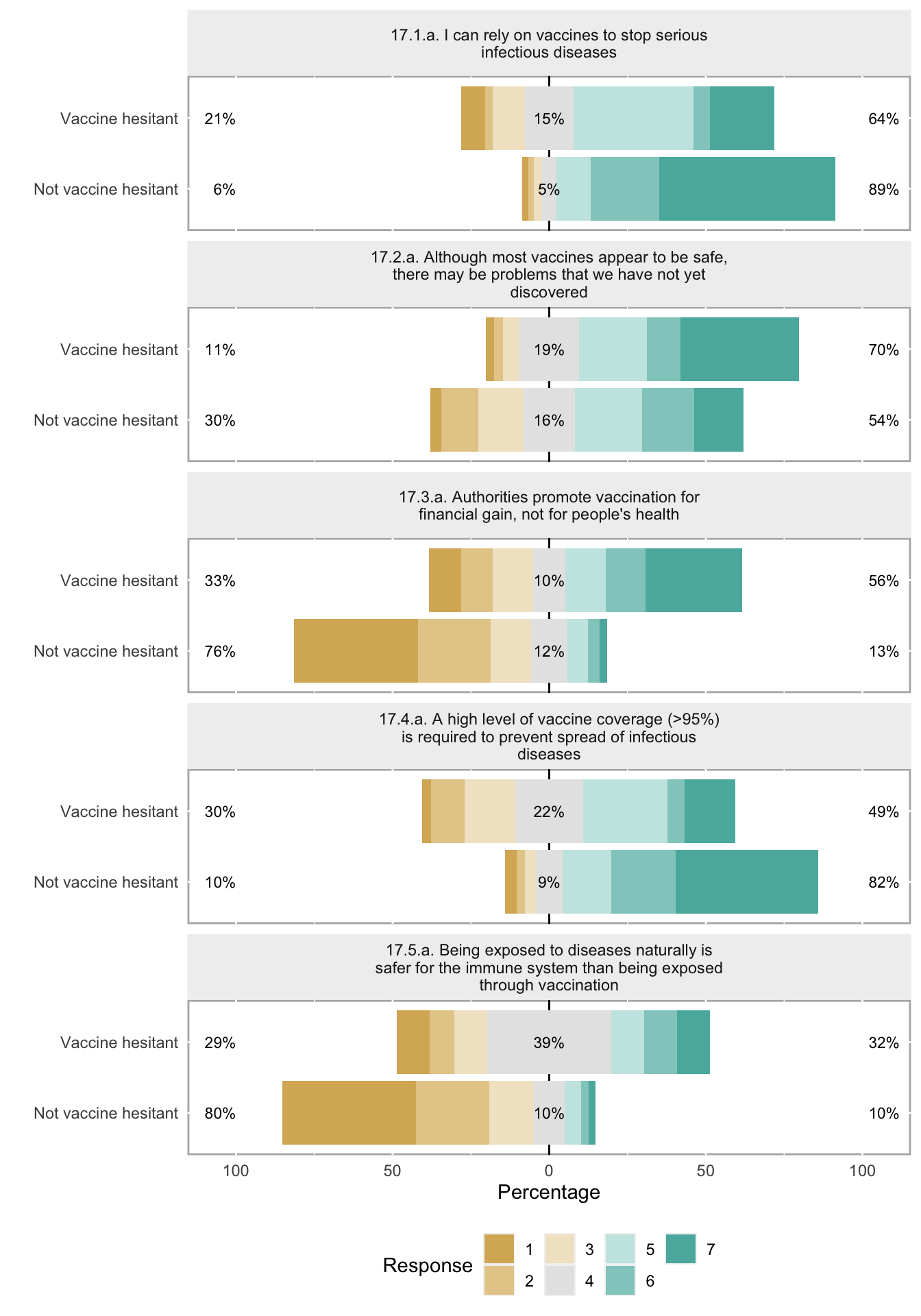

**S2 Figure. Comparison of vaccine confidence scores for vaccine hesitant and non-hesitant participants.** Participants were asked to rank five statements on attitudes to vaccines (the full text of survey question 17 is provided in supplementary information). Ranking was from 1, strongly disagree, to 7, strongly agree. The range of responses for each statement is presented for the hesitant and non-hesitant groups as separate bar graphs with percentages on the left and right indicating disagree (1-3) and agree (5-7) responses, respectively. Note that responses 17.1, 17.2, 17.3 and 17.5 were combined to generate a VAX score.

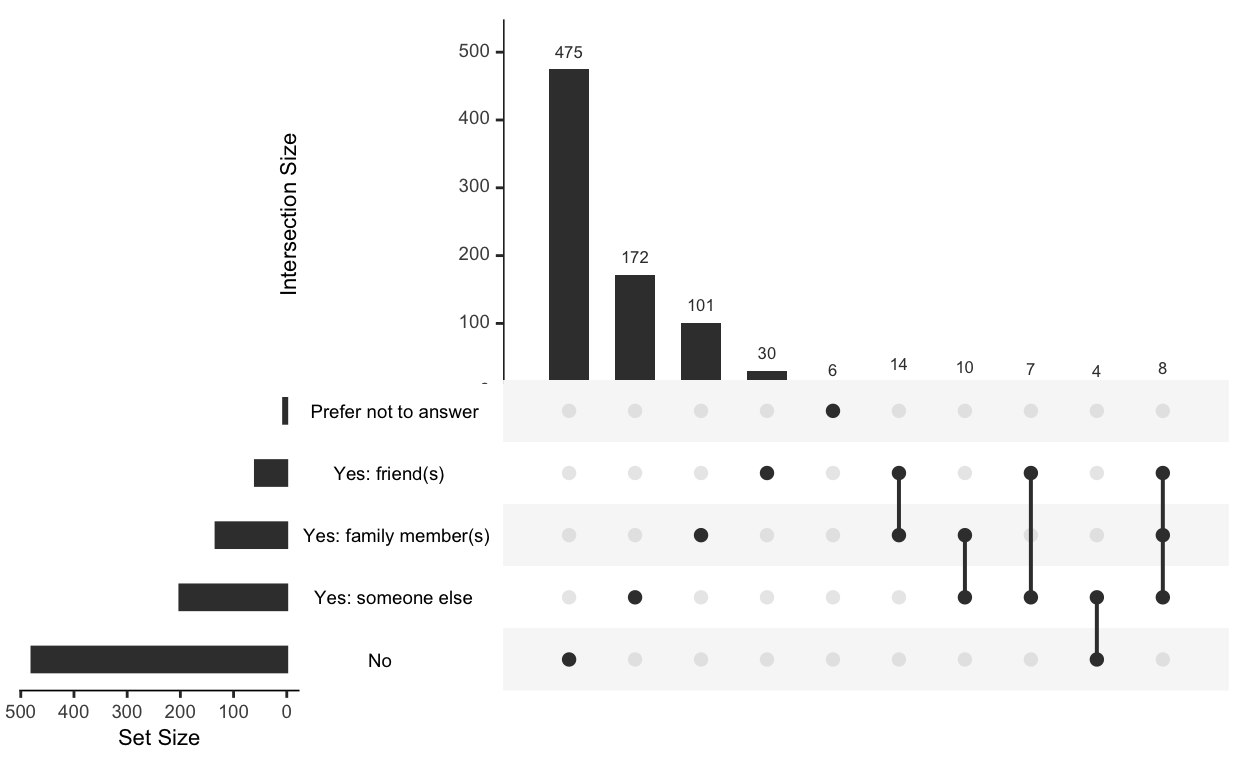

**S3 Figure. Experiences of deaths among friends, family or others.** Question 21 of the survey asked students: “Do you personally know anyone who has died from COVID-19?” Students were allowed to make multiple selections, but in most cases selected one category. Horizontal columns, selection frequency for each individual option; vertical bars, selection frequency for combinations of options.

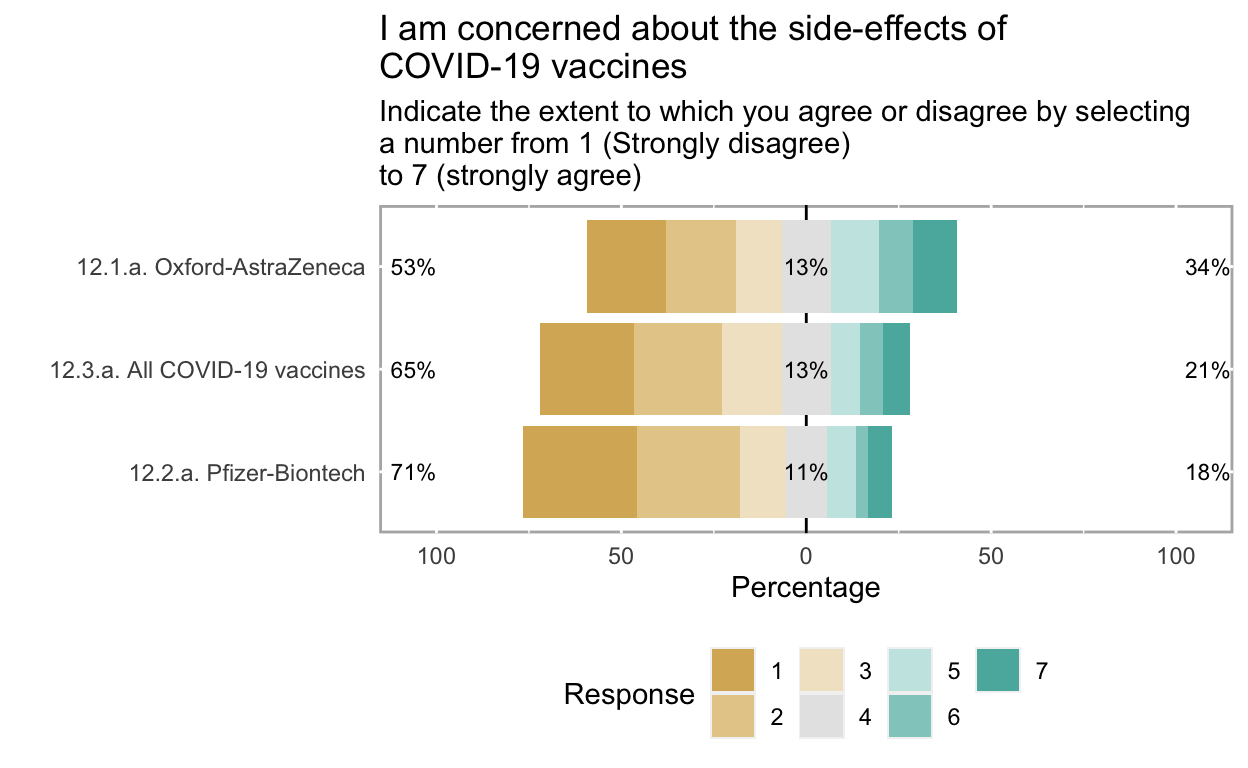

**S4 Figure. Concerns about the side-effects of COVID-19 vaccines**. Question 12 asked participants to indicate the extent to which they agreed with a statement on side-effects (i.e. “I am concerned about the side-effects of COVID-19 vaccines?”) for the Oxford AstraZeneca, Pfizer-BioNTech and all COVID-19 vaccines. The extent of agreement was indicated by selecting numbers from 1 (Strongly disagree) to 7 (strongly agree) or Don’t know. Percentages for each score are shown in a bar graph with combined percentage values for scores of 1-3, 4 and 5-7 being shown on the left, middle and right, respectively.
